# The UT Health Psychological Autopsy Interview Schedule (UTH- PAIS) – Description and Reliability of Diagnoses and Transdiagnostic Personality Measures

**DOI:** 10.1101/2022.04.13.22273746

**Authors:** Thomas D. Meyer, Christopher J. Godfrey, Consuelo Walss-Bass

## Abstract

Few studies have used psychological autopsies to evaluate large and diverse populations on transdiagnostically relevant variables such as personality, temperament, and trauma exposure, rather they tend to focus on specific psychiatric disorders or manner of death. We therefore developed the *UT Health Psychological Autopsy Interview Schedule* (UTH-PAIS). The measure is described, and our results show that that the PAIS diagnoses and dimensions can be reliably assessed. Furthermore, we were able to show that our sample of donated brains overall matches the demographic characteristics of larger pool of individuals receiving a medical autopsy. In the Discussion we review the strengths and potential limitations of the study and outline in which context the PAIS will prove to be useful.

## Introduction

In situations where the circumstances of death are not fully determined or potentially require a forensics investigation, medical examiners use autopsies to determine the cause and manner of death (Choo & Choi, 2012), often including any records and/or reports (e.g., suicide notes) they have access to in the evaluation. However, scant efforts are made to understand the mental state and psychological characteristics of the deceased during these assessments because past psychological assessments (e.g., completed personality inventories) are rarely available.

Ideally, investigators would have access to face-to-face interviews and self-reported questionnaire data about the mental health status, personality, and cognition from individuals before they pass away. However, this level of access is not always possible or practical, especially if standardized assessments, where all individuals complete the same batteries of measures before their death, are required. To overcome this limitation, Psychological Autopsies (PA), which involve in-depth interviews with next-of kin (NOK), have long been used to achieve a better understanding of a decedent’s life, personality, and/or behavior.

Originally developed in the 1960s by Edwin Shneidman (1981) as an approach to study the mental state and motivation of individuals who committed suicide, PA interviews allow researchers to reconstruct a person’s mental well-being during their lifetime. Although Shneidman’s original interviews were performed as an unstructured, case by case idiographic assessment of individuals, which included photos and personal possessions when available, studies have since then used PAs to establish or confirm psychiatric diagnoses and symptoms of mental health disorders in deceased individuals (Cavanagh et al., 2003). Several studies have examined aspects of suicides in different age groups (De Leo et al., 2013; Ross et al., 2017; Sun et al., 2015), or in different cultures (Bhise & Behere, 2016; Chávez-Hernández & Macías-García, 2016; Phillips et al., 2002). By establishing a timeline and potential risk factors for suicide, clinicians gained greater insights into the lives, mental state, and personality of individuals who committed suicide. Given the individual and public health impact of suicides, it is easy to understand why psychological autopsies have primarily focused on this specific topic.

PAs have also been used to better understand the brain mechanisms underlying mental health disorders, by identifying potential differences in brain structures, anatomy, and tissues of clearly defined samples with a specific diagnosis (e.g., dementia, schizophrenia) or symptom clusters when compared with those of healthy controls (e.g. Cairns et al., 2010; Gollub et al., 2013; Innamorati et al., 2008; Knable, 1999; Kövari et al., 2011; Papageorgiou et al., 2016). With this approach studies have shown that there are physiological changes in the brain of those who suffered from mental health disorders such as schizophrenia (Calhoun et al., 2009), alcohol use (Cardenas et al., 2011), or depression (Frodl et al., 2008). Yet another approach was taken by Thompson et al. (2013) who were fortunate enough to have access to both an interview with the individual before their death, and an interview with the individual’s next of kin. This unique opportunity was possible because all patients were already under hospice care at the time of the interview, although being in a hospice care could have also affected the assessments. Additionally, this level of access is not always possible.

While categorical psychiatric diagnoses remain of interest, efforts have recently shifted from diagnostic categories to transdiagnostically relevant features and traits, because of the overlap in symptoms and high rate of comorbidities between mental health conditions (e.g. Aldao et al., 2016; Cerdá et al., 2010; Friborg et al., 2013). To date, these efforts have mostly been confined to suicide with a focus on the completed act, irrespective of the presence or absence of any potential specific psychiatric diagnoses. Ross et al. (2017) examined aspects of personality, recent life events, quality of life, social support, aggression, and impulsivity in individuals who did not have a diagnosable mental health disorder yet committed suicide, compared to persons who died suddenly from other causes. Other studies have examined different aspects of suicide, such as the effect of work stress (Schneider et al., 2011; Shah et al., 2016). Few studies have used PAs to evaluate large and diverse populations on transdiagnostically relevant variables such as personality, temperament, and trauma exposure, rather they tend to focus on specific psychiatric disorders or manner of death. We developed the *UT Health Psychological Autopsy Interview Schedule* (UTH-PAIS) to overcome this gap. While specific psychiatric diagnoses are still assessed, the UTH-PAIS assesses different facets of behavior such as tendency to act impulsively, ability to terminate tasks, resistance to distraction, and self-regulation (Moeller, Barratt, et al., 2001; Moeller, Dougherty, et al., 2001) in order to obtain a comprehensive picture of the personality of an individual during life. Here, we present a detailed description of the UTH-PAIS and its content, including first data about its reliability.

## Methods

### The UT Health Psychological Autopsy Interview Schedule (UTH-PAIS)

The UTHealth Brain Collection (UTHBC) in collaboration with the Harris County Institute of Forensic Sciences (HCIFS), preserves high-quality tissue including brain, blood, and skin fibroblasts, in combination with extensive clinical information to drive evidence-based research. The collection of postmortem tissue is not restricted to individuals who died as the result of suicide or to individuals who have had a specific mental health problem. Rather we collect a) tissues from all individuals whose NOK provided consent for a donation regardless of any specific lifetime history of mental health problems or suicide, and b) whose NOK agree to an interview about the deceased person’s life, well-being, and personality.

A UTH-PAIS is obtained from tissue donors by a phone interview with the NOK 6 to 8 weeks after tissue donation, after informed consent, and with approval from the Institutional Review Board. All interviews are audio-recorded for supervision purposes and training. The UTH-PAIS consists of several parts, described below

1. A brief overview of the decedent’s social status and overall physical health (e.g., physical health issues, marital status, smoking status).
2. Assessment of personality and temperamental traits. In this part of the interview, the NOK is asked to rate the decedent on specific items on a scale from 1 meaning “Not at all” to 10 meaning “Very much”, where 5 represents the average person. These items were selected from validated inventories that attempt to a) reflect the construct in question (e.g., Fun Seeking or Behavioral Inhibition (Carver & White, 1994), Self-regulation (Diehl, 2006; Schwarzer et al., 1999) and b) can be rated based primarily on observations (e.g., verbally aggressive, aggression towards physical objects, and getting involved in physical fights). We also include the personality dimensions which were suggested as the alternative system to categorical personality disorders from the American Psychiatric Association’s Diagnostic and statistical manual of mental disorders (DSM-5; American Psychiatric Association & Association, 2013). These questions assess different aspects of personality, including: Emotional Lability (e.g., “Did [he/she] have unstable emotions, frequent mood changes, or emotions that are intense and out of control?”), Grandiosity (e.g., “Did [he/she] express feelings of superiority, that [he/ she] was entitled to special treatment, and believe that [he/she] was better than others?”), and Cognitive/ Perceptual Aberrations (e.g., Did [he/she] think in odd or unusual ways, see or hears things in unusual or atypical ways, or have speech that was non-detailed or overly detailed, difficult to follow, and frequently going off-topic?). In addition, NOK are asked to rate to what extent these statements applied to the deceased on a scale from 0 “Very little or not at all descriptive” to 3 “Very or Extremely Descriptive”. The NOK is asked to report any psychopharmacological or psychiatric treatment (e.g., ECT, TMS, Light therapy), the decedent might have received during life, including any medications at the time of death.
3. Suicidality is assessed using an adapted version of the Columbia Suicide Severity Rating Scale (Posner et al., 2011).
4. A structured assessment of lifetime history of mental health disorders in line with the DSM-5 (American Psychiatric Association & Association, 2013) is included. The assessments incorporate the criteria for major depressive episodes, (hypo)mania, alcohol use disorder, and substance use disorders, as well as psychotic symptoms. . This is followed by screening questions for panic disorder, agoraphobia, social phobia, specific phobias, generalized anxiety, and obsessive-compulsive disorder (OCD). Unlike the mental health problems mentioned above, a more thorough assessment anxiety disorders and OCD was not intended. The interviewers are therefore instructed to use their clinical judgement, based on DSM-5 criteria, on whether the screening may indicate a clinically relevant problem. An assessment of traumatic life events is conducted based on those listed in the Life Events Checklist for DSM-5 (LEC-5; Weathers et al., 2013). The NOK is asked the age of the decedent at the time of the traumatic event, as well as to rate the negative impact on the decedent’s wellbeing, on a scale from 1 to 10, and if the impact of the events were still ongoing at the time of death.
5. Family history of the deceased, focusing on first-and second-degree relatives, in regards to mental health problems. If a mental health problem is mentioned (including a history of suicide or suicide attempts), it is noted whether any specific mental health problems were formally diagnosed by a healthcare professional, or solely based on the NOKs personal judgement.
6. The Sociodemographic Information section includes questions about the education and professional status of the deceased, including type of employment they had, if any.
7. At the end of the interview, the NOK is provided with information on how to access any bereavement services, or other help if needed. NOK are also asked if they consent to be re-contacted in case additional questions arise, clarifications about the information are needed, and if they consent to be contacted for potential future studies.
8. After the interview is completed, interviewers provide a rating about their judgement of the quality of data provided by the NOK, and whether they feel there was any evidence of social desirability bias based on NOK comments.

### Procedure

The decision was made to conduct the UTH-PAIS over the phone in order to decrease burden to NOKs (e.g., travel time, travel related costs). Up to 3 appointments, if consent is not withdrawn before, are scheduled in case the NOK cannot be reached at the agreed date and time for the UTH-PAIS.

Training of the raters was originally provided by the first author (TDM), a clinically trained and certified psychologist, who was also one of the raters in the majority of the cases.

The training consisted of jointly going through the UTH-PAIS first, and then sitting in on interviews with a trained rater, before being observed doing the interviews till competency is reached according to supervision. If during supervision of the UTH-PAIS, additional questions arise, the interviewer is asked to re-contact the NOK if we have approval to do so. In some cases, more than one NOK will provide information about the deceased individual. For the interrater reliability study, presented here, the assessment was scored by two trained raters independently, either by sitting in on an interview or – as in most cases – by listening to the recordings.

A consensus diagnosis of any psychiatric diagnosis, or designation as a non-psychiatric control, is reached for each individual, according to DSM-5 criteria, after review of all available information, including the UTH-PAIS, any medical records, and the medical examiner’s autopsy report. The diagnosis is determined by a panel of three trained clinicians, including raters involved in the interview, at least one clinical psychologist (TDM), and two psychiatrists.

## Results

### Sample description for interrater reliability

The study sample was comprised of 55 individuals who were between the ages of 19 and 71 at the time of death (M = 47.65, SD = 12.46) and were predominantly male (n = 45, 81.8%). The majority of participants were identified as White (n=43, 78.2%), followed by Black/African American (n=10, 18.2%), and Asian (n=2, 3.6%). Six (10.9%) were identified as Hispanic/Latino, 48 (87.3%) were non-Hispanic/Latino, and 1 (1.8%) were of unknown ethnicity. The majority of the sample was married at the time of death (n=18, 32.7%), some never married (n=16, 29.1%), others were divorced/separated (n=16, 29.1%), in a Domestic

Partnership (n=4, 7.3%), or widowed (n=1, 1.8%). A total of 44 (80%) individuals had completed at least 12 years of education. Nineteen (34.5%) were reported to have been employed full time, 22 (40%) were retired or on disability, with 1 (1.8%) student, and 13 individuals (23.6%) were reported as being unemployed. A significant number of individuals (n=34, 61.8%) were reported to have had a serious medical illness, and 24 (43.6%) had received a diagnosis for major psychiatric conditions at least once. Causes of death were drug toxicity (n=22, 40%), cardiovascular disease (n=21, 38%), respiratory/pulmonary disease (n=3, 5.5), asphyxia (n=3, 5.5), or other (e.g., infection, gunshot wound, or undetermined) (n=6, 10.9%). Eight (14.6%) of all these cases were identified as suicides by the NOK.

The NOK who were interviewed for the study included the decedent’s mother (n=14, 25.4%), with one case where the brother also attended the interview; in four cases the NOKs were the decedent’s father (7.8%) with two of those being step fathers; in 12 cases the NOK was the wife of the deceased (21.8%) and in six cases it was the husband of the deceased (10.9%), with one being a common law husband; in six cases it was the sister (10.9%) and in two cases the brother (3.6%) who provided the relevant information; in three cases the son of the decedent did the interview (5.5%) and in six cases it was the daughter (10.9%). In one of these cases the daughter was joined by the decedent’s ex-wife during the interview; and in two cases close friends of the decedent provided the information (3.6%).

### Interrater reliability results

Table 1 displays the prevalence of assessed mental health problems in this sample, some referring to DSM-5 diagnoses (e.g., bipolar disorder [BD]), and some referring to positive screens for a mental health problem (e.g. social anxiety). Since many individuals had comorbid conditions, there were more diagnoses than cases. Overall rater agreement across categories was very high, and ranged from 78% (Generalized Anxiety Disorder [GAD]) to 98% (Alcohol use disorder [AUD]). Kappa ranged from 0.65 (BD) to 0.96 (AUD), indicating good to excellent reliability, with one outlier being GAD (0.38). Yule coefficients, calculated to account for low base rates in our sample(only 3 cases of BD; Spitznagel & Helzer, 1985), reached the maximum value of 1.00 for several categories, with the lowest score again for GAD (0.63). For four diagnoses we were able to calculate a dimensional score in form of the number of fulfilled criteria and estimated interrater-reliability. The Intraclass-coefficients (ICC) for these dimensions ranged from 0.89 to 0.97, indicating excellent reliability (see Table 1).

**Table 1.** Inter-rater agreement and reliability of Diagnoses and Screening Questions made via the Psychological Autopsy Interview Schedule (UTH-PAIS) (n = 55 interviews)

| Diagnosis | Number of cases | % Agreement | Kappa | Yule coefficient | ICC |
| --- | --- | --- | --- | --- | --- |
| Major Depression Disorder | 17 | .85 | .69 | .73 | .91 |
| Bipolar Disorder | 2 | .96 | .65 | .82 | .89 |
| Psychosis | 5 | .96 | .81 | .88 | ---- |
| Alcohol Use Disorder | 29 | .98 | .96 | 1.00 | .98 |
| Substance Use Disorder | 27 | .95 | .89 | .90 | .97 |
| Panic Attacks<br>*[Screening only]* | 19 | .96 | .92 | 1.00 | ---- |
| Agoraphobia<br>*[Screening only]* | 6 | .93 | .71 | 1.00 | ---- |
| Social Phobia<br>*[Screening only]* | 5 | .96 | .81 | 1.00 | ---- |
| Specific Phobia<br>*[Screening only]* | 11 | .89 | .72 | .80 | ---- |
| Generalized Anxiety Disorder<br>*[Screening only]* | 5 | .78 | .38 | .63 | ---- |
| Obsessive Compulsive Disorder<br>*[Screening only]* | 3 | .96 | .73 | 1.00 | ---- |
| Number of traumata endorsed<br>(at least one)<br>*[Screening only]* | 54 | .98 | n.a.<sup>a</sup> | n.a.<sup>a</sup> | .93 |

*Notes:* Yule Coefficient is a measure that can be informative in addition to kappa when the cell frequencies/prevalences are low; ICC, Intra Class Coefficient, calculated to estimate interrater reliability for dimensional ratings of specific disorders in the form of number of criteria fulfilled.
<sup>a</sup> Kappa could not be calculated when a categorical variable was created for no trauma or at least one trauma endorsed since all individuals reported at least one traumatic event for the deceased person, and there was only one discrepancy between raters.

Interrater-reliability for trait dimensions such as behavior activation, negative urgency, or self-regulation was excellent, with all values obtained between 0.92 and 0.98. When assessing the DSM-5 personality disorder trait domains and facets (Table 2), the ICC ranged from 0.75 for the domain Detachment with six items, to 0.98 for Disinhibition, suggesting good to excellent interrater reliability. Looking at individual facets, the range of ICC across domains was 0.55 for Restricted Affectivity to 0.96 for Deceitfulness, suggesting moderate to excellent reliability depending on the facet. Switching to the internal consistency for the domains, Cronbach’s α ranged from 0.65 for Disinhibition to 0.96 for Negative Affectivity. The lower reliability of the original DSM-5 Disinhibition scale seemed to be related to the poor performance of the reverse coded facet ‘Rigid Perfectionism’ showing even a negative corrected item-total correlation (Table 2). When eliminating this facet as done by Suzuki et al. (2015), the internal consistency increased to 0.84, with all four corrected item-total correlations being adequate between 0.60 and 0.70.

**Table 2.** Inter-rater reliability for Personality traits and personality disorder facets and domains.

| **Personality Scale** | **ICC** | **Personality domains and their facets** | **Alpha (Cronbach)** | **ICC** | **M (SD)** | **r<sub>it</sub>** |
| --- | --- | --- | --- | --- | --- | --- |
| Behavior Activation Scale (3 items) | .96 | **Antagonism** | .84 | .95 | 0.98 (0.87) |  |
| Behavior Inhibition Scale (3 Items) | .93 | Manipulativeness |  | .90 | 1.00 (1.29) | .54 |
|  |  | Deceitfulness |  | .96 | 0.80 (1.19) | .76 |
|  |  | Grandiosity |  | .76 | 1.04 (1.31) | .81 |
|  |  | Attention Seeking |  | .87 | 0.96 (1.10) | .43 |
|  |  | Callousness |  | .92 | 0.76 (0.97) | .65 |
|  |  | Hostility |  | .79 | 1.32 (1.15) | .49 |
| Impulsivity (1 item) | .94 | **Disinhibition <sup>a</sup>** | .65 | .98 | 1.28 (0.78) |  |
| Introversion (3 items) | .92 | Irresponsibility |  | .94 | 1.00 (1.23) | .67 |
|  |  | Impulsivity |  | .90 | 1.36 (1.25) | .68 |
|  |  | Distractibility |  | .92 | 0.84 (0.99) | .58 |
|  |  | Risk taking |  | .67 | 1.32 (1.35) | .52 |
|  |  | Rigid Perfectionism (-) |  | .71 | 1.88 (1.17) | -.21 |
| Negative Urgency (1 item) | .98 | **Negative Affectivity** | .86 | .85 | 1.42 (0.69) |  |
| Perseverance (2 items) | .96 | Emotional Liability |  | .82 | 1.52 (1.16) | .84 |
|  |  | Anxiousness Separation |  | .82 | 1.09 (1.13) | .59 |
|  |  | insecurity |  | .82 | 1.26 (1.21) | .79 |
|  |  | Submissiveness |  | .89 | 0.65 (0.96) | .60 |
|  |  | Hostility |  | .79 | 1.13 (1.06) | .58 |
|  |  | Perseveration |  | .73 | 1.13 (1.14) | .36 |
|  |  | Depressively |  | .93 | 1.44 (1.24) | .75 |
|  |  | Suspiciousness |  | .72 | 1.04 (1.11) | .61 |
|  |  | Restricted Affectivity (-) |  | .55 | 2.35 (1.03) | .14 |

**Table 2 (continued)**
| **Personality Scale** | **ICC** | **Personality domains and their facets** | **Alpha (Cronbach)** | **ICC** | **M (SD)** | **r<sub>it</sub>** |
| --- | --- | --- | --- | --- | --- | --- |
| Self-Regulation (2 items) | .98 | **Psychoticism** | .83 | .86 | 0.73 (0.93) |  |
| Verbal & Physical Aggression (3 items) | .82 | Unusual Beliefs |  | .82 | 0.84 (1.14) | .78 |
|  |  | Eccentricity |  | .75 | 0.72 (0.94) | .68 |
|  |  | Cognitive & Perceptual Aberrations |  | .80 | 0.64 (1.04) | .82 |
| Well-being Scale (4 items) | .96 | **Detachment** | .68 | .75 | 1.01 (0.69) |  |
|  |  | Withdrawal |  | .63 | 0.96 (1.17) | .64 |
|  |  | Intimacy Avoidance |  | .54 | 0.76 (1.13) | .69 |
|  |  | Anhedonia |  | .70 | 0.64 (1.22) | .48 |
|  |  | Depressively |  | .93 | 1.36 (1.22) | .25 |
|  |  | Restricted Affectivity |  | .55 | 0.68 (1.10) | .26 |
|  |  | Suspiciousness |  | .72 | 1.04 (1.10) | .23 |

*Notes:* The personality ratings are based on 55 cases, while the analyses on personality disorder facets and domains rely on ratings for 25 deceased individuals since these ratings were added to the UTH-PAIS at a later stage;
<sup>a</sup> When eliminating the recoded item ‘Rigid Perfectionism’, the Domain Disinhibition scale mean dropped to 1.13 (SD = 1.00), with ICC=.87; its psychometrics improved with Cronbach’s alpha = .84, and all corrected item-total correlations ranging between .60 to .70.

### Is the brain donation sample representative of all medical autopsies?

We were able to compare some variables with the publicly available data from the Harris County Institute of Forensic Sciences (HCIFS) for 2016-2019. When comparing the two samples, the frequencies of causes of death of our total sample and those of the HCIFS did not significantly differ (χ^2^ (4, n=18812) = 8.20, p =.09), with for example, 12.9% and 11.1% respectively determined to be suicides. The main observed difference was in homicide cases, with 1.9% and 9.9% respectively. The gender ratio of general cases undergoing a medical autopsy and of cases where we received a brain donation did not differ significantly, 27.3% females in the UTHBC versus 23% in the IFS (χ^2^ (1, n=18831) = 1.04, n.s.). Exploring race, and differentiating because of cell frequencies only between White, Black, Asian, and Other (e.g., Native American, unspecified), there was a highly significant difference between the two samples, (χ^2^ (3, n=14478) = 480, 24, p < .001). The percentages of White (58% and 62.8%) and Asian (5% and 3.5%) individuals were similar, but the IFS had more medical autopsies involving Black/African American individuals (36.1%) than the UTBC (20.4%). The opposite was the case for the remaining categories that includes either Native American (IFS: n = 10, UTHBC: n = 0) or undetermined race with 0.3% and 13.3% respectively. The ratio of Hispanics vs. Non-Hispanic individuals did not differ, with 23.4% and 20%, respectively (χ^2^ (1, n=18732) = 0.41, n.s.). In regards to age groups (< 18, 18-39, 40-64, >64), the UTHBC received fewer donations from minors and individuals 65 years and older, and slightly more cases in the middle age categories (χ^2^ (3, n=18722) = 18.22, p < .01).

## Discussion

The present study describes the rationale and methodology for a new Psychological Autopsy Interview Schedule developed at the University of Texas Health Science Center at Houston (UTH-PAIS). This interview schedule was designed with the goal of being able to assess not only traditional mental health problems such as mood disorders in deceased individuals, but also to identify transdiagnostically relevant factors such personality factors, temperament, and traumatic experiences in these individuals. In addition to providing an overview over the UTH-PAIS, we also present data on the reliability of the assessed categorical and dimensional variables. Overall, the results demonstrate good to excellent inter-rater reliability after training. We also demonstrate that most derived dimensional measures (e.g., Extraversion-Introversion, Negative Affectivity, or number of fulfilled criteria for alcohol use disorder) have sufficient internal consistency to be reliable for further analyses. The one exception was the personality domain Disinhibition, due to lack of correlation of the reverse coded Rigid Perfectionism facet as mentioned above. However, a review of literature shows there is discrepancy as to whether this facet should be considered part of the Disinhibition domain or not (e.g., Krueger et al., 2012; Suzuki et al., 2015).

Because some inherent factors linked to medical autopsies, such as the involvement of forensic investigations, could bias the type of donations a postmortem tissue collection receives, we compared the donations received by the UTHBC to the general yearly composition of the samples undergoing a medical autopsy at the HCIFS, based on the public reports from the same time period. Overall, the UTHBC sample was comparable to the HCIFS in regards to gender, ethnicity, and cause of death. We found differences related to race and age distribution. The UTHBC received more donations from individuals between 18 and 64 years of age, than from younger or older people. In regards to the latter, the discrepancy may be related to neurological conditions, which are not the focus of our research and are not explicitly assessed in the UTH-PAIS. In the case of minors, there might a hesitation of parents to the brain donation. The few individuals under the age of 18 who’s parents agreed to the donation were all mid to late adolescence. As far as race, the UTHBC received approximately 16% fewer donations if the decedent was identified as Black or African American, and proportionally more donations from Caucasian and those whose race was not definitely determined. Future research is needed to determine the generalizability of this data to other less urban and/or culturally distinct populations.

We do not have specific information as to why the NOK choose not to consent to brain donation. The surrogate donors are likely grieving their loved one’s death at the time they are approached about donations (on average within 24 h of death), which could make the decision difficult. In addition, the viewpoint of the surrogate may involve cultural or spiritual differences and perspectives in regards to tissue donation in general, or there may be concerns related to the genetic and biological research attached to the donation. Therefore, future research is needed to determine attitudes and beliefs surrounding postmortem brain donations.

Reliability is a necessary condition for validity but not a guarantee. Ideally, the postmortem diagnoses and personality ratings would be validated against medical and psychological reports obtained during the lifetime of the decedent, but this kind of data is rarely available. While some studies have compared personality ratings of individuals and relatives and found significant agreement in ratings (e.g., Thompson et al., 2013), it would of importance to explore the convergent validity between an individual’s subjective responses and a close relative or loved one for the current interview schedule. It might also be helpful to explore, if feasible, whether the quality of interview data differs depending on the relationship of the informant to the deceased one (e.g., mother, father siblings, and spouses).

There are some additional limitations to the study. Because the death of a loved one is a very difficult time emotionally, we felt it was not ethically justified to conduct the interview a second time by a different interviewer in order to assess interrater reliability. We therefore relied on either the second rater sitting in on the original telephone interview, or listening to the recording. In the former case, the two rater’s cannot see each other’s ratings. However, it is possible that observing the interview conducted by someone else could increase the likelihood of the listening rater gaining insight about the other person’s rating. In these cases, the observed interrater-reliability estimates must be considered as reflecting the upper limits of reliability, after training under ideal conditions. However, given the good to excellent reliability of most ratings, we believe this is not a major concern for our study.

In summary, to date only a few studies have attempted to use psychological autopsies to evaluate such a large and diverse population without focusing solely on specific disorders or manner of death. We were able to show that the UTH-PAIS can be considered a reliable instrument not just to assess traditional psychiatric disorders but also to assess transdiagnostically relevant variables such as personality, temperament, and trauma exposure. Future studies will assess the association between these dimensional measures of personality, socioeconomic indicators, and brain-derived biological measures we obtain from the UTHBC. The availability of the information obtained from the UTH-PAIS, together with biological tissues from individual subjects represents an unprecedented opportunity to advance the understanding of how disruption of biological mechanisms and brain circuits can modulate behavior.

## Data Availability

All data produced in the present study are available upon reasonable request to the authors.

## Acknowledgements

We are grateful to Nic Crist, Nikki LaRosa, Priel Meir, and Katherine Najera who have greatly helped with the interview process and entering the data.

